# Elexacaftor/Tezacaftor/Ivacaftor Reshapes Airway Inflammation and Proteomic Landscape in Cystic Fibrosis

**DOI:** 10.1101/2025.10.02.25337177

**Authors:** Filip Årman, Stefanie Diemer, Lotta J. Happonen, Lisa I. Påhlman

**Author notes:** Equal contribution (Lotta J. Happonen, Lisa I. Påhlman). Corresponding author: Lisa I. Påhlman.

## Abstract

**Background:** Elexacaftor/Tezacaftor/Ivacaftor (ETI) has significantly improved clinical outcomes for people with cystic fibrosis (pwCF), but the molecular effects on airway inflammation remains incompletely understood. This study aimed to characterise longitudinal changes in airway inflammation and sputum proteomes following ETI treatment, and to correlate proteomic shifts to changes in inflammatory cytokines.

**Patients and methods:** Sputum from pwCF (*n*=30) were collected before start of ETI and after 3 and 9-12 months of treatment. Sputum from healthy control subjects (*n*=7) were included for comparison. Samples were analysed for total proteome content using data independent acquisition liquid chromatography tandem mass spectrometry (DIA LC-MS/MS), and inflammatory cytokines using Mesoscale assays. Protein expression trends were analysed using k-means clustering, and correlations between airway proteomes and inflammatory cytokines were performed using Pearson correlation and enrichment analysis.

**Results:** ETI therapy resulted in significant changes in the airway proteome, mainly related to decreased neutrophil degranulation and an increase in anti-proteases. Levels of IL-1*β*, IL-8, and TNF*α* decreased with ETI therapy, which correlated with proteins involved in neutrophil degranulation. In contrast, IL-6 levels increased and correlated with proteins involved in O-glycosylation of mucins. Despite these improvements, proteomic and cytokine profiles remained distinct from healthy controls after 9-12 months.

**Conclusion:** ETI leads to broad shifts in airway protein expression in pwCF with reduced neutrophilic inflammation and restored protease/antiprotease balance. Despite these changes, there is still increased airway inflammation compared to healthy control sputum. This dataset provides a valuable resource for further exploration of CF airway biology under ETI therapy.

## Background

Cystic fibrosis (CF) is a genetic disorder caused by mutations in the *cystic fibrosis transmembrane conductance regulator* (*CFTR*) gene, leading to impaired transport of chloride (Cl^-^) and bicarbonate (HCO3-) ions across epithelial surfaces (1). In the airways, loss or dysfunction of CFTR results in dehydrated, viscous mucus and defective mucociliary clearance. This altered microenvironment facilitates microbial colonization, driving persistent bacterial infections, sustained neutrophilic inflammation, and progressive decline in lung function (2). In recent years, CFTR modulators that directly improve the expression and function of the CFTR channel have transformed CF care. In December 2022, the triple combination therapy Elexacaftor/Tezacaftor/Ivacaftor (ETI) was approved in Sweden for the treatment of CF. ETI combines CFTR “correctors”, which improve protein folding and trafficking to the cell surface, with a “potentiator” that enhances channel gating (3). This dual action increases both the surface expression and functional activity of CFTR, restoring chloride and fluid transport. ETI has led to significant improvements in lung function, exacerbation rates, and quality of life for people with CF (pwCF) (4, 5). Moreover, ETI therapy has been associated with reduced systemic (6–9) and airway inflammation (10–12).However, despite these clinical improvements, many individuals remain persistently infected with CF pathogens such as *Pseudomonas aeruginosa* (13), indicating that airway infection and inflammation may persist even under ETI therapy. To optimize treatment strategies, it is essential to elucidate how ETI reshapes airway pathophysiology at the molecular level.

Mass spectrometry-based proteomics of sputum samples offers a powerful, unbiased approach to investigate the complex proteome-level dynamics shaping airway disease. Unlike more targeted assays, label free DIA LC-MS/MS enables comprehensive profiling of molecular signatures and dysregulated biological pathways, increasing our understanding of the pathophysiology behind airway disease. Importantly, integrating proteomic data with selected cytokine profiles provides a systems-level view of airway biology, linking global proteome changes to inflammatory processes.

In this study, we employed DIA LC-MS/MS to characterize the airway proteome in sputum from pwCF collected before and after ETI initiation, alongside healthy controls. Additionally, we quantified targeted cytokines using Mesoscale assays. By characterizing both proteomic and cytokine changes, we defined proteomic alterations associated with CF and ETI therapy and established a framework for biomarker discovery and mapping of biological pathways that may guide future precision medicine approaches. Notably, we were able to quantify a broader range of proteins in CF sputum post-ETI compared to earlier studies, revealing previously uncharacterized biological mechanisms and cytokine–proteome correlations.

## Methods

### Study population

Adult pwCF attending the CF centre at Skåne University Hospital, Lund, Sweden, were eligible for inclusion. Exclusion criteria were age <18 years, ongoing treatment with triple modulator therapy, and inability to expectorate sputum at baseline. Sputum samples were collected at regular visits to the clinic prior to start of ETI treatment and after 3 and 9-12 months of treatment, respectively. The participants had to be clinically stable without signs of ongoing exacerbation at the time of sampling. Healthy individuals without known pulmonary conditions or anti-inflammatory treatment were recruited as controls.

Clinical data were collected from the medical journal. Lung function evaluation, including Forced expiratory volume in 1 second in percent of predicted (FEV1pp), was performed at the time of sputum sampling. Chronic *P. aeruginosa* infection was defined according to Leeds criteria (14).

The study was approved by the Swedish Ethical Review Authority (reference numbers 2021-01191 and 2024-00361-01). Written informed consent was obtained from all participants.

### Sputum sampling and preparation

PwCF donated expectorated or induced sputum using tailored hypertonic saline. Healthy controls donated an induced sputum using 8% NaCl. All sputum collection was supervised by a physiotherapist. Sputum samples were homogenized using 0.1% dithiothreitol (DTT) (Sigma-Aldrich) as described before (15). After centrifugation at 1000×g for 10 min, the cell-free supernatant was collected and stored at −80°C until analyses.

### Quantification of inflammatory cytokines

Sputum concentrations of IL-1*β*, IL-6, IL-8 and TNF*α* were quantified using Mesoscale immunoassays (Mesoscale Diagnostics LLA., Maryland, USA) as described previously (16). If cytokines were undetectable, the value was set to the lowest limit of detection (0.1 pg/mL).

### Statistical analyses

Statistical analyses of lung function and inflammatory cytokines were performed using GraphPad Prism 10.2.3 software (GraphPad Software, San Diego, CA). Paired observations of FEV1pp were analysed using paired T-test. Mann-Whitney U test was used for the comparison of cytokine concentrations between groups. The limit for statistical significance was set to 5% (p<0.05).

### Sample preparation for mass spectrometry (MS)

For MS analysis, 10 μl of DTT-treated, cell-free, sputum samples prepared as described above were digested to peptides as described (17). After reconstitution, 20 μl of each sample was loaded onto Evotip C18 trap columns (Evosep Biosystems) according to the manufacturer’s instructions.

### Liquid chromatography tandem mass spectrometry (LC-MS/MS)

The samples were analysed using a 30 samples per day method (Evosep Biosystems, connected to PepSep MAX C18 15cm x 150μm, 1.5μm column, Bruker Daltonics) and data collected a timsTOF HT (Bruker Daltonics) mass spectrometer with a diaPASEF method, combining data-independent-acquisition (DIA) of spectra with parallel accumulation-serial fragmentation (PASEF) (18).

### MS database search

The database search was carried out using the software DIA-NN 1.8.1 (Data-Independent Acquisition by Neural Networks) in library free mode using the reviewed *Homo sapiens* proteome (UP000005640 release 2024_10) as a database. The precursor m/z was set to 250-1450, and fragment m/z to 200-1700. The precursor charge was set to 2-4, with a minimum peptide length of 7-47 amino acids. Trypsin was set as the enzyme, with cleaving after prolines not included and a maximum number of missed cleavages of 2. The false-discovery rate (FDR) was filtered at precursor level at 1 % (0.01).

### MS data analysis

All MS data analysis steps were made with the DPKS v0.1.5 (19) python package if nothing else is stated. Raw precursor values were median normalized and Log2 transformed using the normalize method in DPKS. The normalized precursor values were quantified to proteins with the DPKS quantify method using a combined approach, where the top five most abundant precursors per protein were applied to the maxLFQ algorithm for extracting the relative quantification signal across all analysed samples.

Unipressed 1.3.0 (20) package was used to retrieve protein to gene annotations from UniprotKB on 2025-01-27. For the generation of heatmaps, the quantified protein matrix was filtered from NA values and scaled on protein level using the DPKS scale method with zscore setting, and plotted with PyComplexHeatmap 1.7.6 (21). For differential expression analysis, a two-sided t-test was applied using the DPKS compare method allowing for a minimum four valid sample values per group and using Benjamini-Hochberg to correct for multiple testing, controlling at a FDR of 5% (0.05). Gene enrichment analyses were performed using Metascape (22). For K-means clustering of, we used scikit-learn (23) v1.1.2 KMeans, and the optimal number of clusters was investigated applying the elbow method on 30 simulated clusters.

### Preprocessing of inflammatory cytokine data for correlation analysis

Prior to logarithmic transformation, a pseudocount of 1 was added to all raw quantified values, and the data was Log2 scaled to make the data adhere to a normal distribution.

### Correlation analysis

For each cytokine, the mean expression for the samples across all three timepoints were calculated. Cytokine trends were tested for correlations against all MS protein mean expression trends in R 4.4.2 (24) using cor.test function with the Pearson method.

### Functional enrichment of cytokine correlated proteins

Proteins that had a top 25% percentile correlation or anti-correlation to their corresponding cytokines were considered for downstream enrichment analysis. Each set of cytokine-associated proteins were enriched using the ReactomePA (25) enrich Pathway function with all identified proteins as a background, adjusting for 0.5 (50%) p-value cutoff and 0.9 (90%) q-value cutoff to compensate for a small background set of proteins and large search space in the Reactome database.

## Results

### Patient characteristics

A total of 30 pwCF and seven healthy controls were included in the study (**Supplementary figure 1**). All pwCF provided at least one follow-up sample, and 22 participants (73%) contributed sputum at both follow-up time points (**Supplementary figure 1**, highlighted in bold). Three patients did not provide sputum at 9-12 months due to inability to expectorate (n=1) or discontinuation of ETI treatment due to adverse effects. Baseline data for the study participants are presented in **Table 1**. Notably, male gender was dominating in the CF group (77%) but underrepresented among controls (29%), and 57% of pwCF were receiving lumacaftor/ivacaftor therapy prior to start of ETI. Consistent with previous reports (4), ETI treatment resulted in a significant improvement in lung function, with mean FEV1pp increasing from 67.7% (SD 19.4) at baseline to 83.1% (SD=17.7, p<0.001) at three months, and 82.7% (SD=20.5, p<0.001) at 9-12 months (**Supplementary figure 2**).

**Table 1.** Patient characteristics at baseline.

| Demographic and clinical data | CF (n=30) | Controls (n=7) |
| --- | --- | --- |
| Age years, median (IQR) | 37 (29–46) | 46 (32–62) |
| Male gender n (%) | 23 (77) | 2 (29) |
| Homozygous $\Delta F508$ n (%) | 19 (63) | NA |
| FEV1% pred. median (IQR) | 64 (56–83) | NA |
| Chronic *P. aeruginosa* n (%) | 16 (53) | NA |
| Pancreatic insufficiency n (%) | 25 (83) | NA |
| Treatment with lumacaftor/ivacaftor n (%) | 17 (57) | NA |

### Changes in the sputum proteome following ETI treatment

To assess the impact of ETI on airway inflammation at the proteomic level, sputum samples from pwCF and controls were analysed using DIA LC-MS/MS. Across all samples, 4952 unique human proteins were quantified, each identified with at least one proteotypic peptide. Heatmap visualization of overall protein expression revealed distinct proteomic profiles at baseline, 3 months on ETI, 9-12 months on ETI, and in healthy controls (**Supplementary figure 3**).

Differential expression analysis using two-sided t-test revealed significant proteomic shifts following ETI initiation. At 3 months, 105 proteins were significantly downregulated and 102 significantly upregulated compared to baseline (**Figure 1A**). Downregulated proteins included neutrophil granular proteins such as neutrophil collagenase (MMP8), bactericidal permeability-increasing protein (BPI) and protein S100-A9. Upregulated proteins included Fc-receptor like protein 5 (SCGB3A1), clusterin (CLU), uteroglobin (SCGB1A1), and anti-proteases such as alpha-1 antitrypsin (SERPINA1) and antileukoproteinase (SLPI) (**Figure 1A**). Gene enrichment analyses indicated that the downregulated proteins were predominantly associated with neutrophil degradation and proteasome pathways, while upregulated proteins were linked to extracellular matrix-related pathways and salivary secretion (**Supplementary Figure 4A-B**).

**Figure 1.**
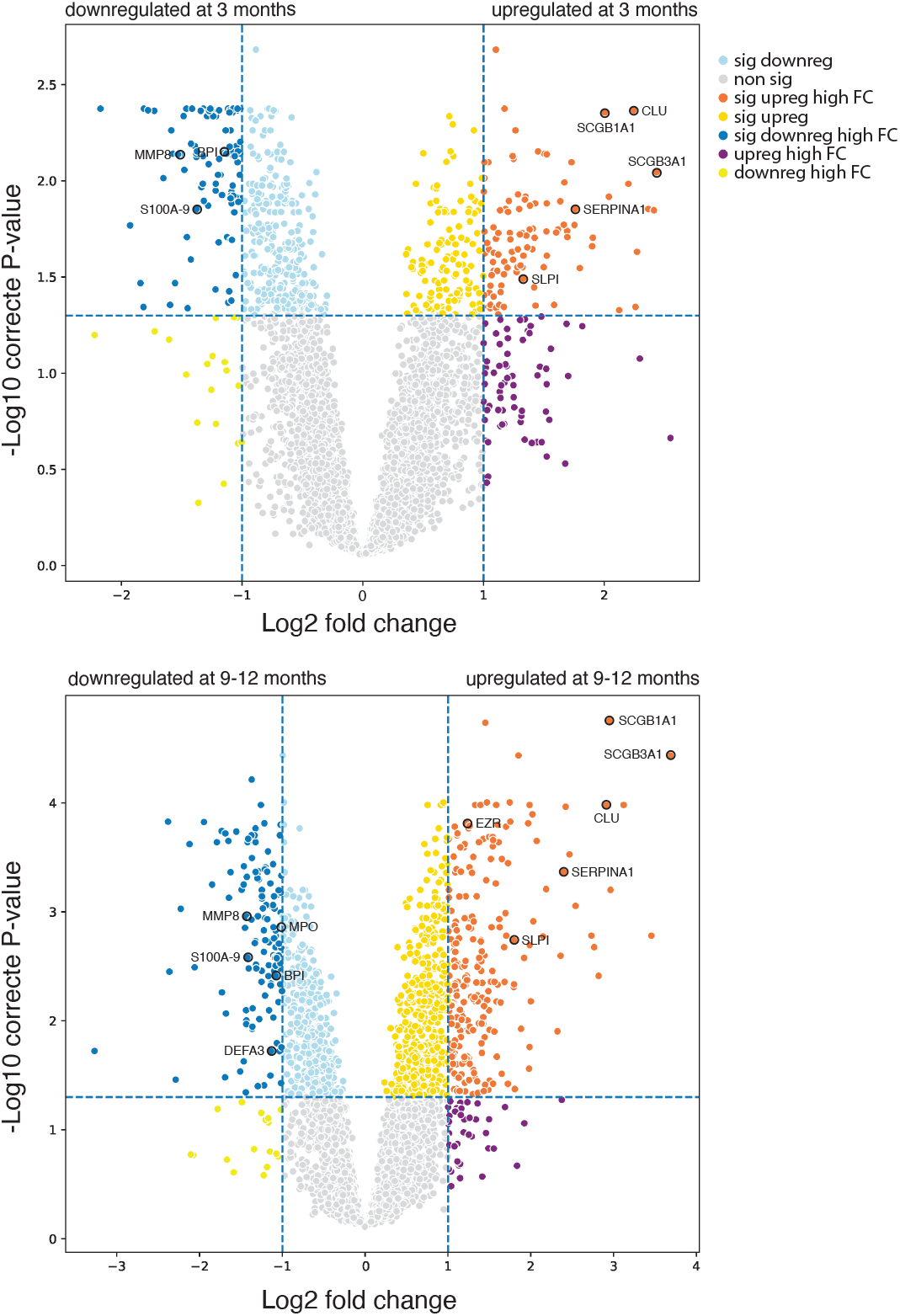
Volcano plot analysis of protein expression differences. The volcano plots depict the differentially expressed proteins after **A)** 3 months and **B)** 9-12 months of ETI treatment compared to before start of ETI. Significantly upregulated proteins (p < 0.05) with a high fold change (FC) (Log2 FC > 1) are shown in orange, significantly downregulated proteins (p < 0.05) with a high FC in dark blue, upregulated proteins meeting the significance threshold but not the FC are shown in dark yellow, downregulated proteins meeting the significance threshold but not the FC are shown in light blue, upregulated proteins meeting the FC threshold but not the significance level are shown in dark purple, downregulated proteins meeting the FC threshold but not the significance level are shown in light-yellow, and finally non-significantly regulated proteins are shown in light grey. Selected proteins of interest are highlighted.

At 9-12 months, 133 proteins were significantly downregulated and 235 significantly upregulated compared to baseline (**Figure 1B**), indicating continued dynamic shift in the proteome composition with prolonged ETI treatment. Notably, additional proteins of interest were downregulated at 9-12 months, including myeloperoxidase (MPO) and neutrophil defensin 3 (DEFA3), while *e*.*g*. ezrin (EZR) was upregulated. These findings support previous reports of upregulation of antileukoproteinase and ezrin, as well as downregulation of protein S100A-9 following ETI treatment (10, 12). Enrichment analyses of up- and downregulated proteins showed similar pathway involvement as at 3 months (**Supplementary figure 4 C-D**). Noteworthy, no significant differences were observed between sputum proteomes at 3 and 9-12 months (**Supplementary figure 5A**). This is consistent with the lung function data where the significant increase in FEV1%pred was stabilized between 3 and 9-12 months (**Supplementary figure 2**). Despite the clearly observable changes in proteome composition after ETI initiation, pwCF proteomes remained distinct from healthy controls even after 9-12 months on ETI (**Supplementary figure 5B**).

To explore longitudinal protein expression trajectories during ETI treatment, we generated 18 K-means clusters of protein expression trends based on the elbow method on 30 simulated clusters (**Supplementary figures 6-7**). In response to ETI, protein expression patterns increased in some clusters (clusters 5, 11, 12, 13, 15 and 17), decreased in others (cluster 0, 2, 4, 6, 10, 9 and 14), while several remained stable or showed inconsistent changes (**Supplementary figure 7**). Clusters with decreasing expression were enriched for proteins involved in *e*.*g*. immune system responses and neutrophil degranulation (clusters 4 and 14), including MMP8, BPI, MPO and DEFA3. Clusters with increasing protein expression included complement-related proteins (cluster 5), EZR, SERPINA1 and SLPI (clusters 11 and 12) (**Figure 2**). Functional annotation showed that clusters with downregulated protein expression during ETI were associated with *e*.*g*. neutrophil degranulation, intracellular protein transport, regulation of vesicle-mediated transport and vesicle organization pathways, whereas clusters with downregulated protein expression were dominated by processes involved in supramolecular fibre and intermediate filament organization, haemostasis, complement and coagulation cascades, salivary secretion and varying signalling pathways, to list a few (**Supplementary figure 8**).

**Figure 2.**
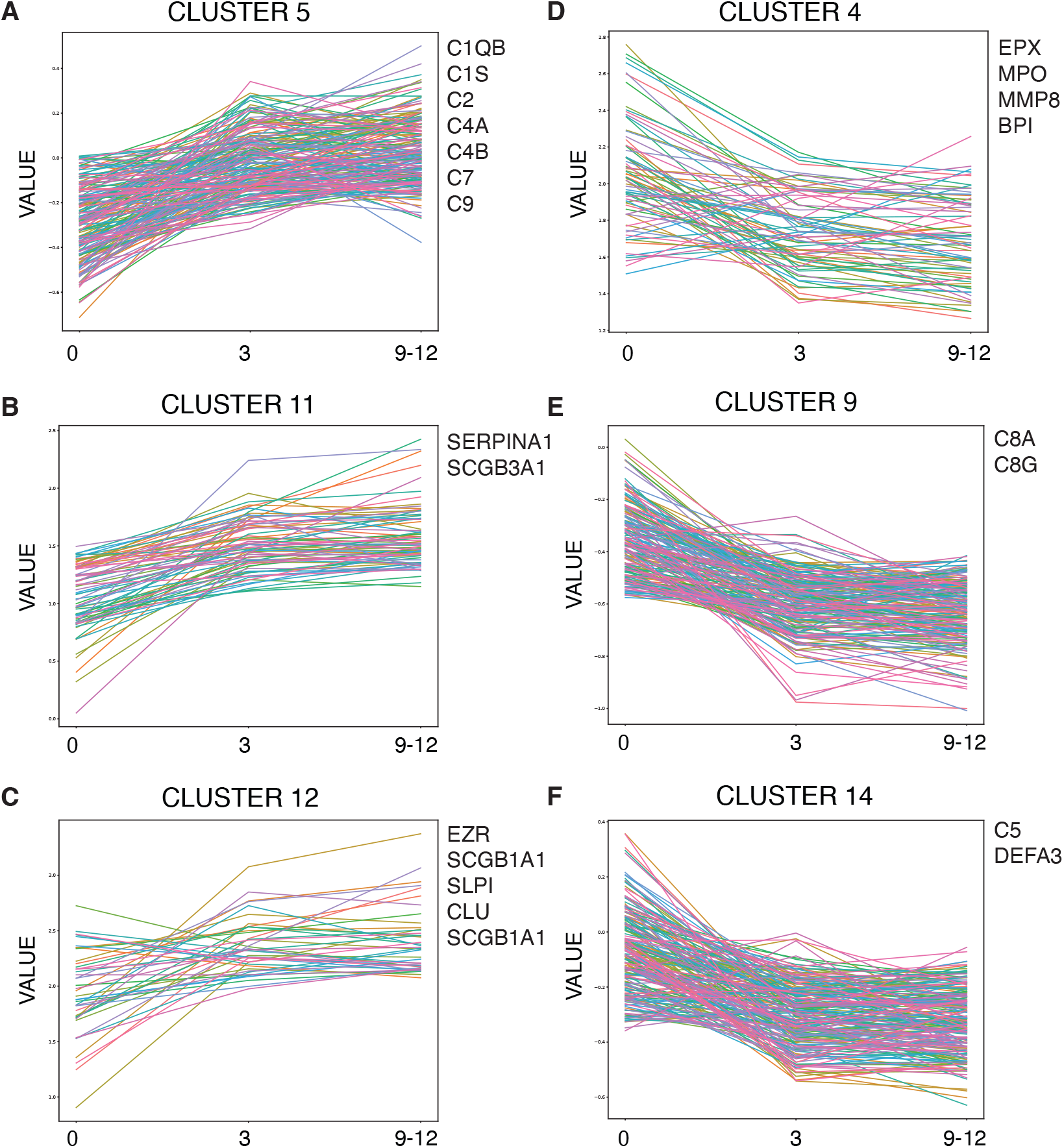
K-means cluster analysis. Cluster analysis of changes in protein expression patterns during ETI treatment. Selected clusters of interest including **A-C)** upregulated proteins and **D-F)** downregulated proteins are shown. Associated proteins of importance are listed next to each cluster.

### Associations between sputum cytokine levels and the airway proteome in response to ETI

To complement the proteomic data, we next quantified inflammatory cytokines in sputum using Mesoscale assays, targeting low-abundant proteins not captured in the total proteome analysis. Following ETI initiation, a significant decline was detected in IL-1*β*, IL-8 and TNF*α* levels (**Figure 3A-C**), while IL-6 concentrations increased (**Figure 3D**). Despite ETI therapy, sputum levels of all analysed cytokines remained elevated in pwCF at 9-12 months compared to healthy controls.

**Figure 3.**
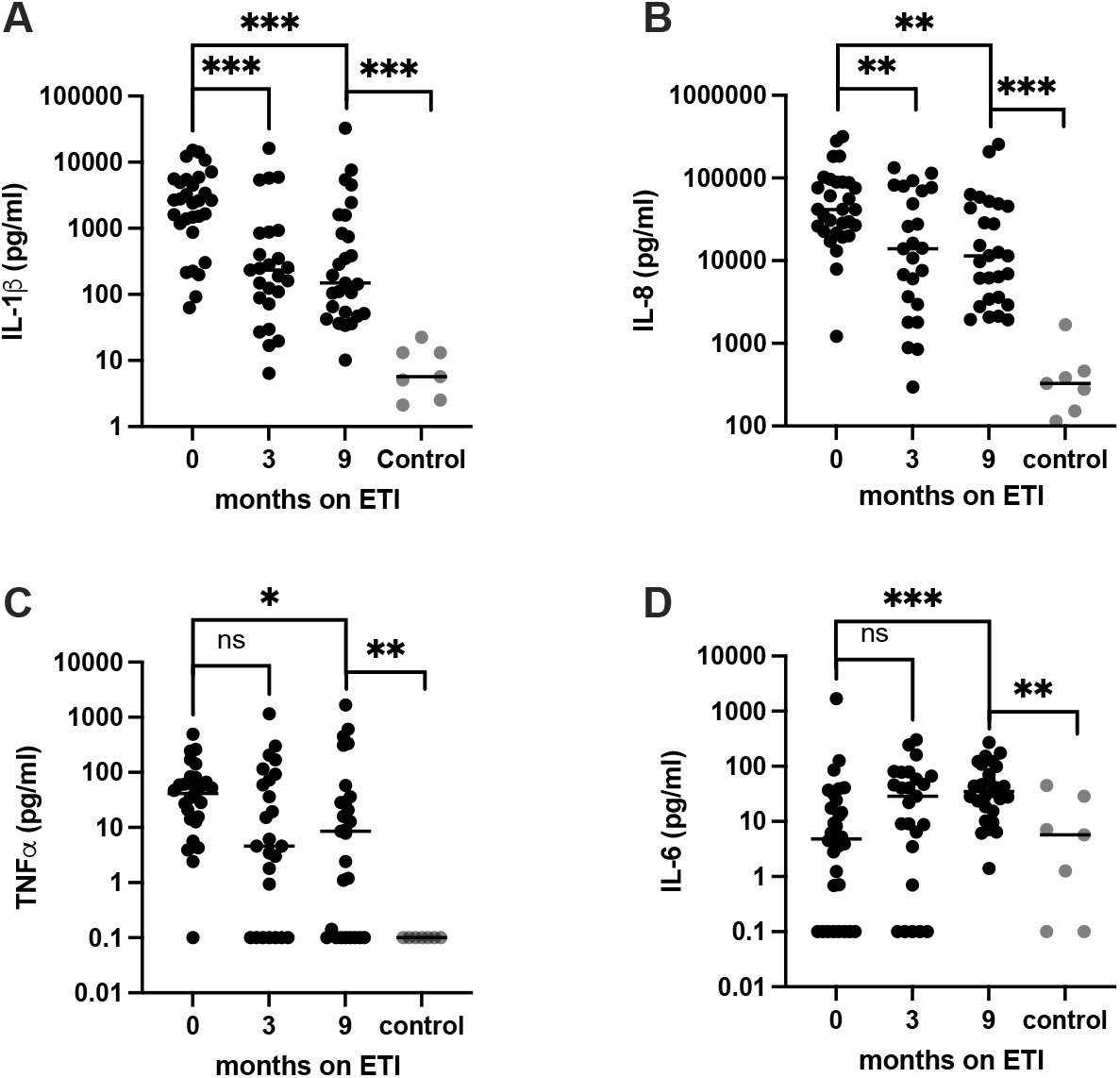
Sputum concentrations of inflammatory cytokines. Sputum samples from pwCF (black dots) and healthy controls (grey dots) were analysed for **A)** IL-1*β*, **B)** IL-8, **C)** TNF*α* and **D)**, IL-6 using Mesoscale assays. The bars represent median values. *=p<0.05, **=p<0.01, ***=p<0.001. ns=non-significant.

To further explore the relationship between cytokine dynamics and proteomic changes following ETI initiation, we performed Pearson correlation analysis (**Figure 4, Supplementary figure 9**). A positive correlation was observed between decreasing IL-1*β* levels during ETI treatment and proteins involved in neutrophil degranulation and DNA replication, including proteins such as neutrophil elastase (ELANE), MMP8, BPI, protein S100A-9 and histones (**Figure 4A**). Similar correlations were found for TNF*α* (**Figure 4B**). In contrast, increasing IL-6 levels during ETI treatment were positively associated with proteins involved in O-linked glycosylation (**Figure 4C**). No significant correlations were identified for IL-8.

**Figure 4.**
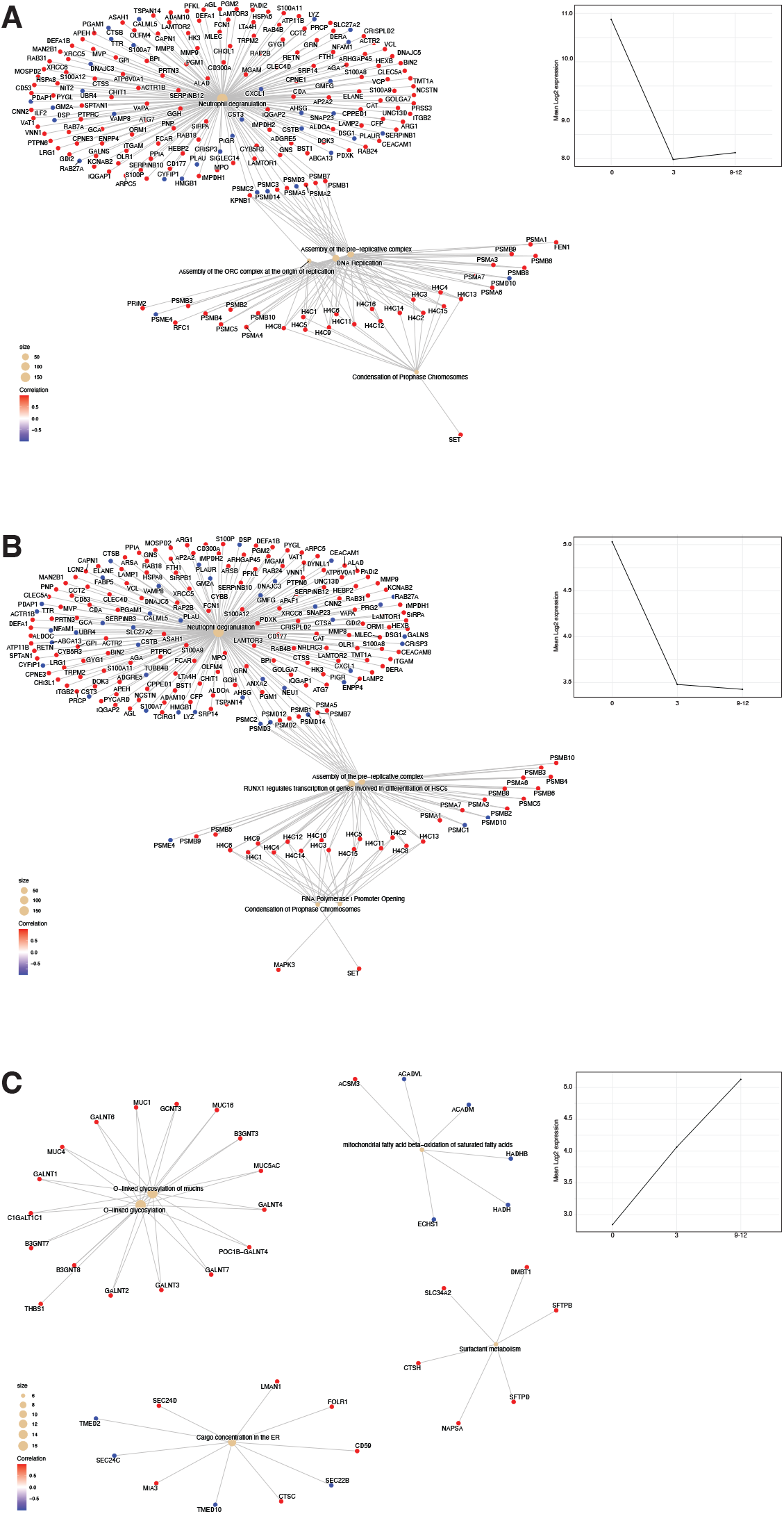
Systemic biological processes associated to cytokine expression after ETI treatment. The network graph shows reactome pathway-enriched protein expressions correlated with the mean cytokine expression data for A) **IL-1**b, **B)** TNF-a and **C)** IL-6 with the line graph for each displayed in the line graph to the left.

## Discussion

The introduction of ETI has markedly improved lung function and quality of life for pwCF. In this study, we provide a comprehensive longitudinal mapping of sputum proteome dynamics and airway inflammation in pwCF before and after ETI initiation. Our findings demonstrate a consistent downregulation of neutrophil-associated proteins and pathways, including MPO, collagenase, BPI, S100A9, and defensins, reflecting reduced neutrophilic inflammation (**Figure 1**). These changes are consistent with previous reports (10) and align with clinical observations of improved lung function and fewer exacerbations following ETI initiation (26). In parallel with the observed reduction in neutrophilic inflammation, we detected increased levels of the key anti-proteases SERPINA1 and SLPI. These proteins are essential for maintaining the protease/anti-protease balance that is critical for pulmonary homeostasis (27). Consistent with prior studies (6), our findings suggest that ETI treatment contributes to restoring this balance.

Among the most significantly upregulated proteins were SCGB3A1 and SCGB1A1, both previously linked to ETI response (10). While the function of SCGB3A1 remains unclear, <u>SCGB1A1</u> is known to exert anti-inflammatory effects by regulating monocyte-induced inflammation (28). Additional upregulated proteins included complement components, CLU and EZR. Elevated levels of complement factors may reflect decreased consumption due to reduced complement activation. Notably, the complement component C5, which in its active form serves as a potent chemoattractant for neutrophils (29), decreased following ETI treatment, in contrast to many other complement components. Increased expression of CLU, a complement inhibitor that protects against (30), and EZR, which regulates airway inflammation by linking CFTR to TLR4 (31, 32), further support a shift toward reduced inflammation.

Sputum concentrations of IL-1β, IL-8, and TNF decreased following ETI treatment, whereas IL-6 levels, which were lower in pwCF than in healthy control sputum at baseline, increased after start of ETI treatment. Reduced IL-6 levels in sputum have previously been reported in pwCF but not in other pulmonary diseases, suggesting that suppressed airway IL-6 expression may be a unique feature of CF-related inflammation (33, 34). Although the underlying mechanisms remain incompletely understood, one proposed explanation is the proteolytic degradation of IL-6 by serine proteases (35). Thus, the observed increase in IL-6 may reflect reduced protease activity due to improved protease/anti-protease balance under ETI therapy.

The decline in sputum IL-1β and TNF*α* following ETI treatment correlated with proteins involved in neutrophil degranulation and DNA replication pathways (**Figure 4**). Notably, proteins associated with DNA replication were predominantly histones, which are released during formation of neutrophil extracellular traps (NETs). In CF, NET formation has been associated with neutrophilic inflammation and reduced lung function (36–38). IL-6 dynamics were associated with increased mucin O-glycosylation (**Figure 4C**), a process central to airway immune regulation. Altered mucin glycosylation is increasingly recognized in chronic lung diseases and may influence mucin immunomodulatory properties (39). Further studies are warranted to explore how ETI affects mucin glycosylation and its functional consequences.

A key strength of this study is the integration of cytokine profiling with DIA LC-MS/MS-based proteomics, allowing for the in-depth evaluation of how inflammatory cytokines are associated with proteome profiles in sputum. We quantified nearly 5000 individual proteins, which is a significant increase in the current data repository for sputum proteome level changes during ETI treatment (10–12). This high number of quantified sputum proteins, combined with this size of a longitudinal cohort, enables a more detailed analyses of biological pathways and their associations. Notably, all enrolled participants provided at least one follow-up sputum, ensuring a CF cohort representative of varying disease severity. Thus, the study provides a rich dataset for future investigations into airway biology and ETI response.

Limitations include the single-centre design and small cohort size, which may affect generalizability. The most severely ill patients were already started on ETI via compassionate use programs and therefore not enrolled in the study. Additionally, ETI therapy led to reduced sputum volumes. These changes in volume are difficult to assess and were not adjusted for in our analyses.

In conclusion, ETI treatment leads to reduced neutrophilic inflammation, improved protease/anti-protease balance, and a significant remodelling of the airway proteome in pwCF. Despite these improvements, proteomic and cytokine profiles remain distinct from those of healthy controls, highlighting persistent airway inflammation and the need for individualized therapeutic approaches. The dataset generated in this study offers a valuable resource for further investigations into airway protein interactions and CF pathophysiology under ETI therapy.

## Data Availability

All proteomic data will become available online upon acceptance of the pre-print.

## Acknowledgements

The authors wish to thank the physiotherapists at the CF centre at Skåne University Hospital Lund, Sweden, for invaluable help with sputum collection, and Malgorzata Berlikowski at Division of Infection Medicine, Lund University, Sweden, for excellent technical assistance. Support from the Swedish National Infrastructure for Biological Mass Spectrometry (BioMS) is gratefully acknowledged. This study was funded by the Swedish Government Funds for Clinical Research (ALF, grant 40204), the Swedish Heart and Lung foundation (grant F2022/2227), the Alfred Österlund Foundation (grant F2022/190), the Swedish Cystic Fibrosis Association (grant F2022/625), the Swedish Research Council (grant 81248 to LIP and 2022-03860 to LJH), the Science for Life Laboratory (SciLifeLab) PALS program, and the Knut and Alice Wallenberg Foundation, the Faculty of Medicine at Lund University and Region Skåne (WCMM Lund, grant 81234).

