## Supplemental figures for "Elexacaftor/Tezacaftor/Ivacaftor Reshapes Airway Inflammation and Proteomic Landscape in Cystic Fibrosis"

### SUPPLEMENTARY FIGURES

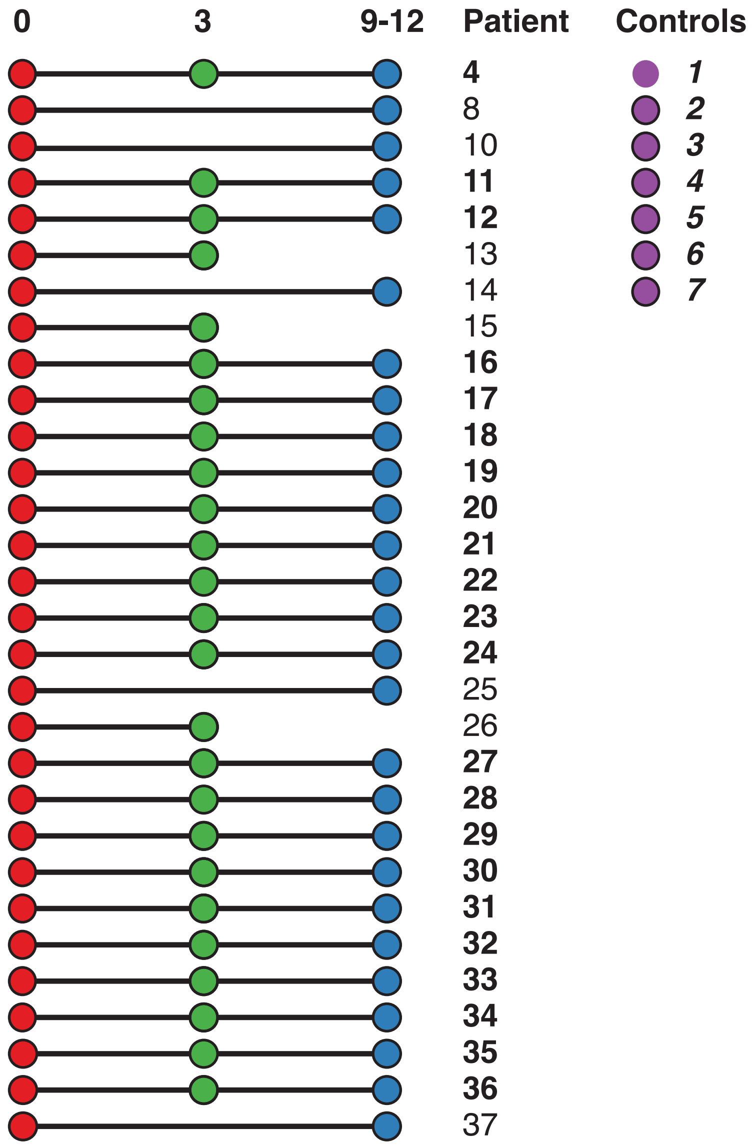


**Supplementary figure 1. Overview of sputum samples included in the study.** The figure indicates the samples collected from each pwCF at a given timepoint, and the samples collected from healthy controls. Samples collected at 0 months are shown as red spheres, at 3 months as green spheres, at 9-12 months as blue spheres and the healthy control group as purple spheres.

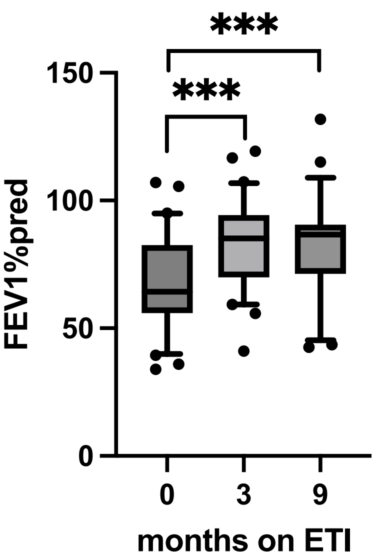


**Supplementary figure 2. Lung function (FEV1pp).** The box plots show FEV1pp in pwCF (*n*=30) before ETI initiation and after 3 and 9-12 months of treatment. ***=p<0.001.

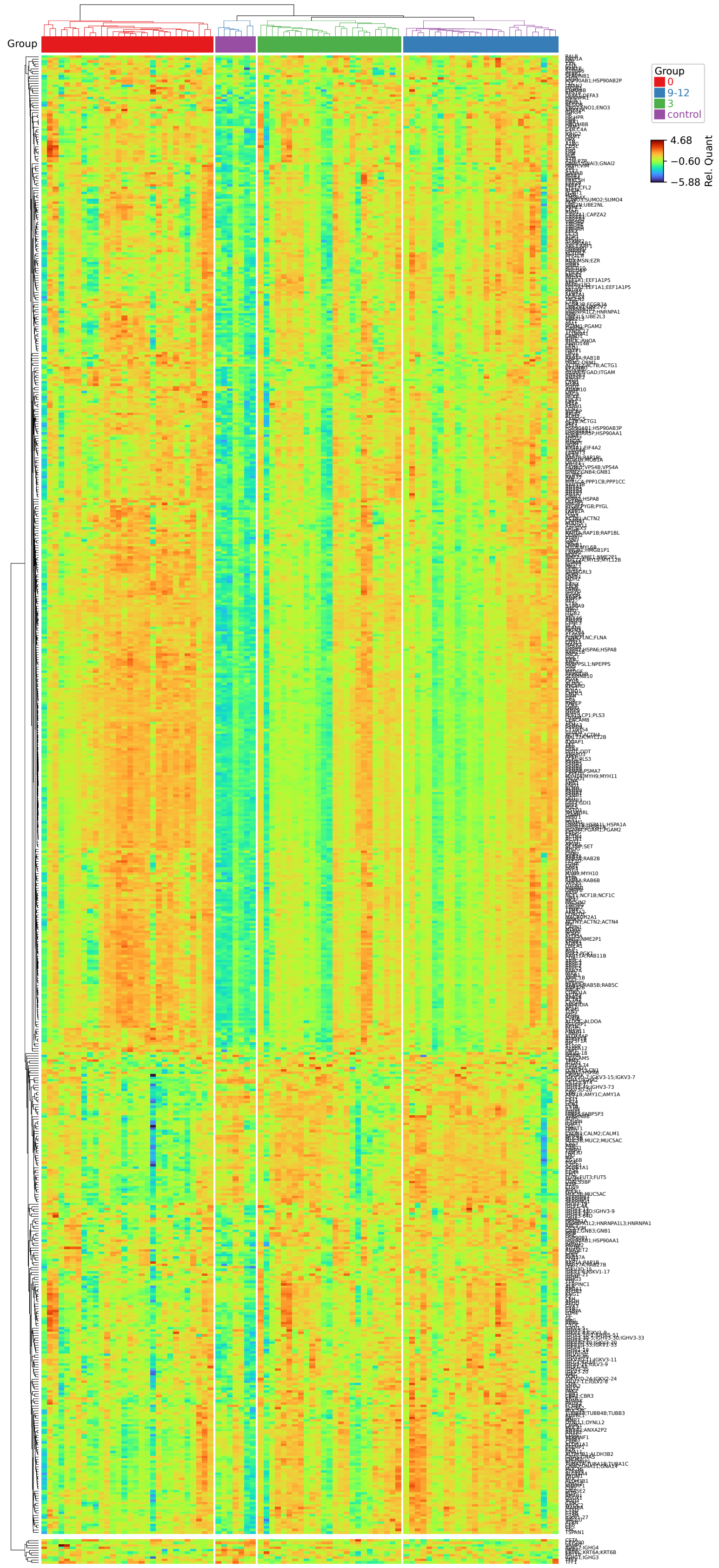


**Supplementary figure 3. Heatmap of overall protein expression profiles.** Log2 transferred and row-wise z-scored MS proteins, grouped samples collected from pwCFs at 0 months are shown in red, at 3 months in green, at 9-12 months in blue and the healthy control group in purple.

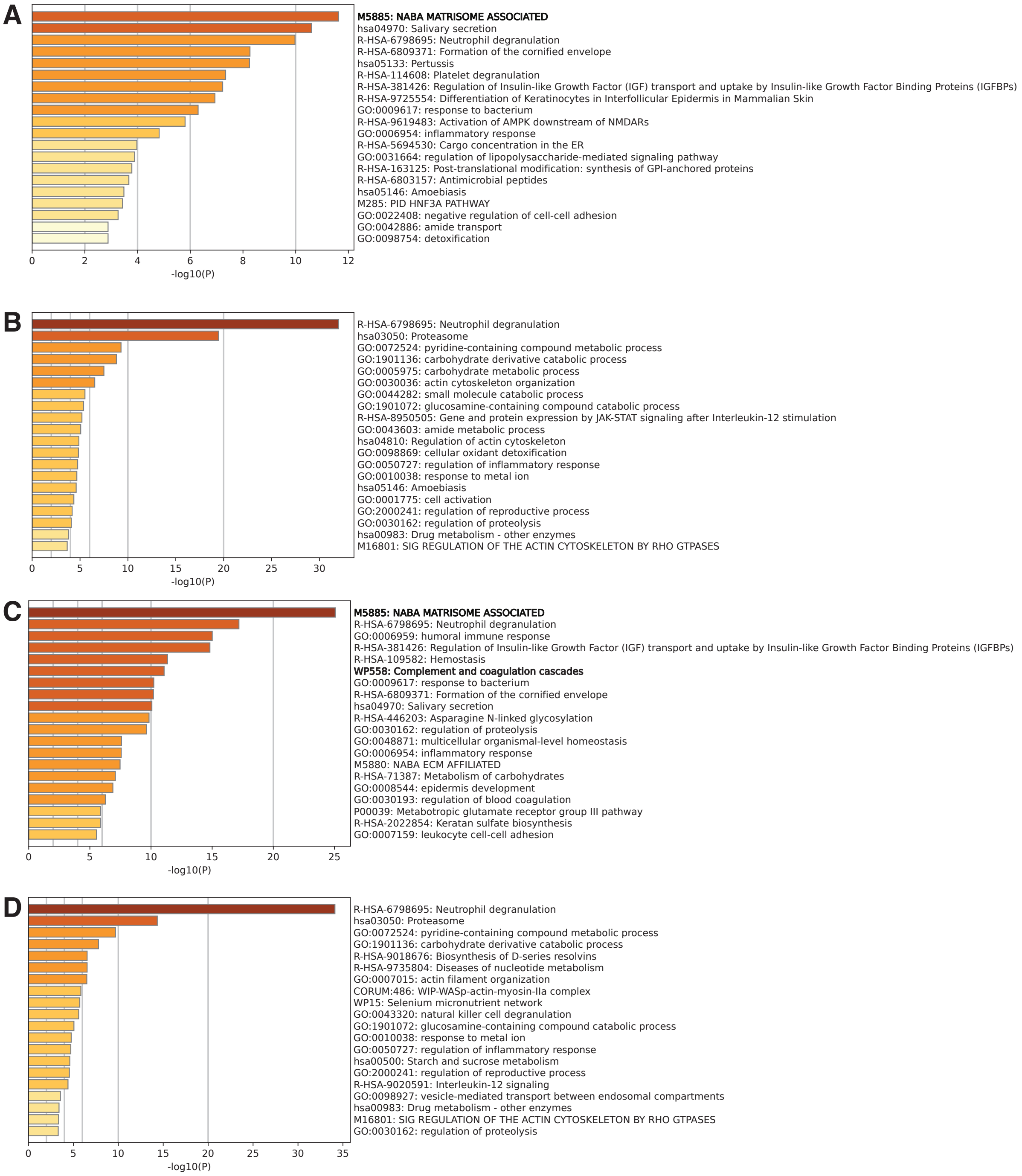


**Supplementary figure 4. Metascape analysis of differentially expressed proteins.** The metascape analysis reveals the functional enrichment of the **A)** significantly upregulated proteins and **B)** significantly downregulated proteins ($p < 0.05$) with a high fold change (FC) (Log2 FC > 1) after 3 months of ET1 treatment, as well as **C)** significantly upregulated proteins and **D)** significantly downregulated proteins ($p < 0.05$) with a high fold change (FC) (Log2 FC > 1) after 9-12 months of ET1 treatment. Selected groups of enriched proteins are highlighted in bold font.

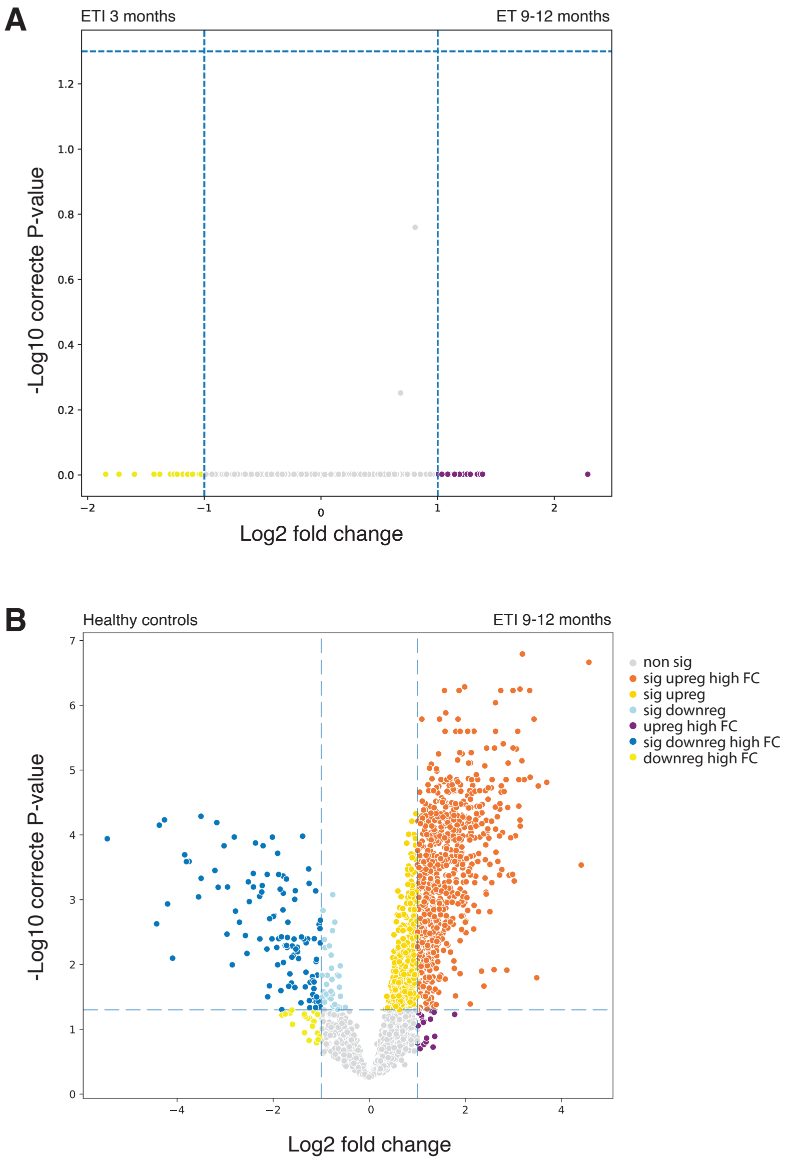


**Supplementary figure 5. Volcano plot analysis of protein expression differences.** The volcano plots depict the differentially expressed proteins after **A)** three months compared to 9-12 months of ETI treatment, and **B)** 9-12 months after ETI treatment compared to healthy controls. Significantly upregulated proteins ($p < 0.05$) with a high fold change (FC) (Log2 FC> 1) are shown in orange, significantly downregulated proteins ($p < 0.05$) with a high FC in dark blue, upregulated proteins meeting the significance threshold but not the FC are shown in dark yellow, downregulated proteins meeting the significance threshold but not the FC are shown in light blue, upregulated proteins meeting the FC threshold but not the significance level are shown in dark purple, downregulated proteins meeting the FC threshold but not the significance level are shown in light-yellow, and finally. non-significantly regulated proteins are shown in light grey. Selected proteins of interest are highlighted.

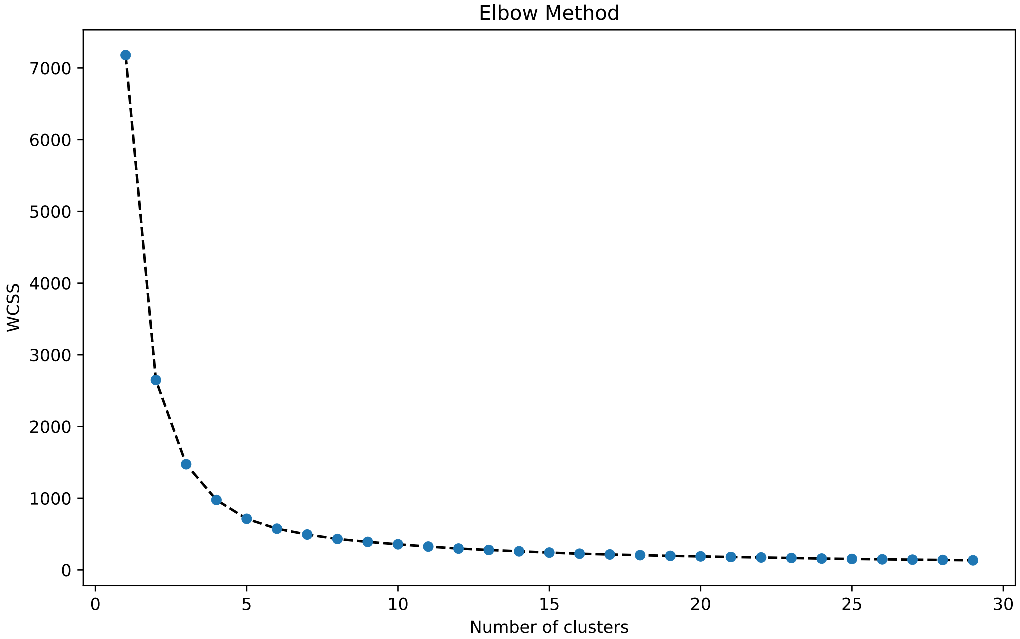


**Supplementary figure 6. K-means cluster analysis.** The elbow plot shows Within-Cluster Sum of Squares (WCSS) against the number of k-means clusters sampled for the dataset.

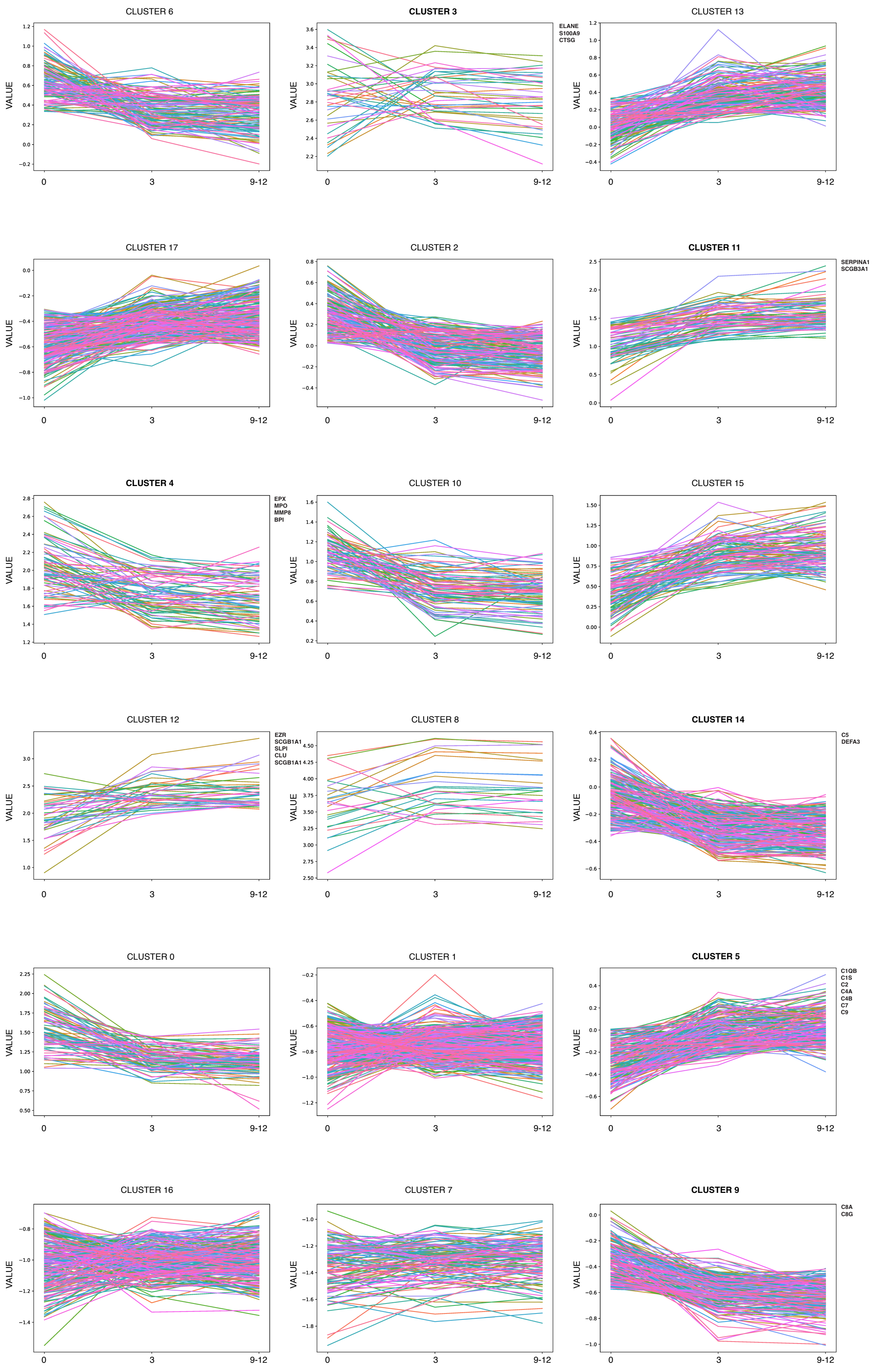


**Supplementary figure 7. K-means cluster analysis of changes in protein expression patterns during ETI treatment showing all 18 generated clusters.** Associated proteins of importance is listed next to each cluster.

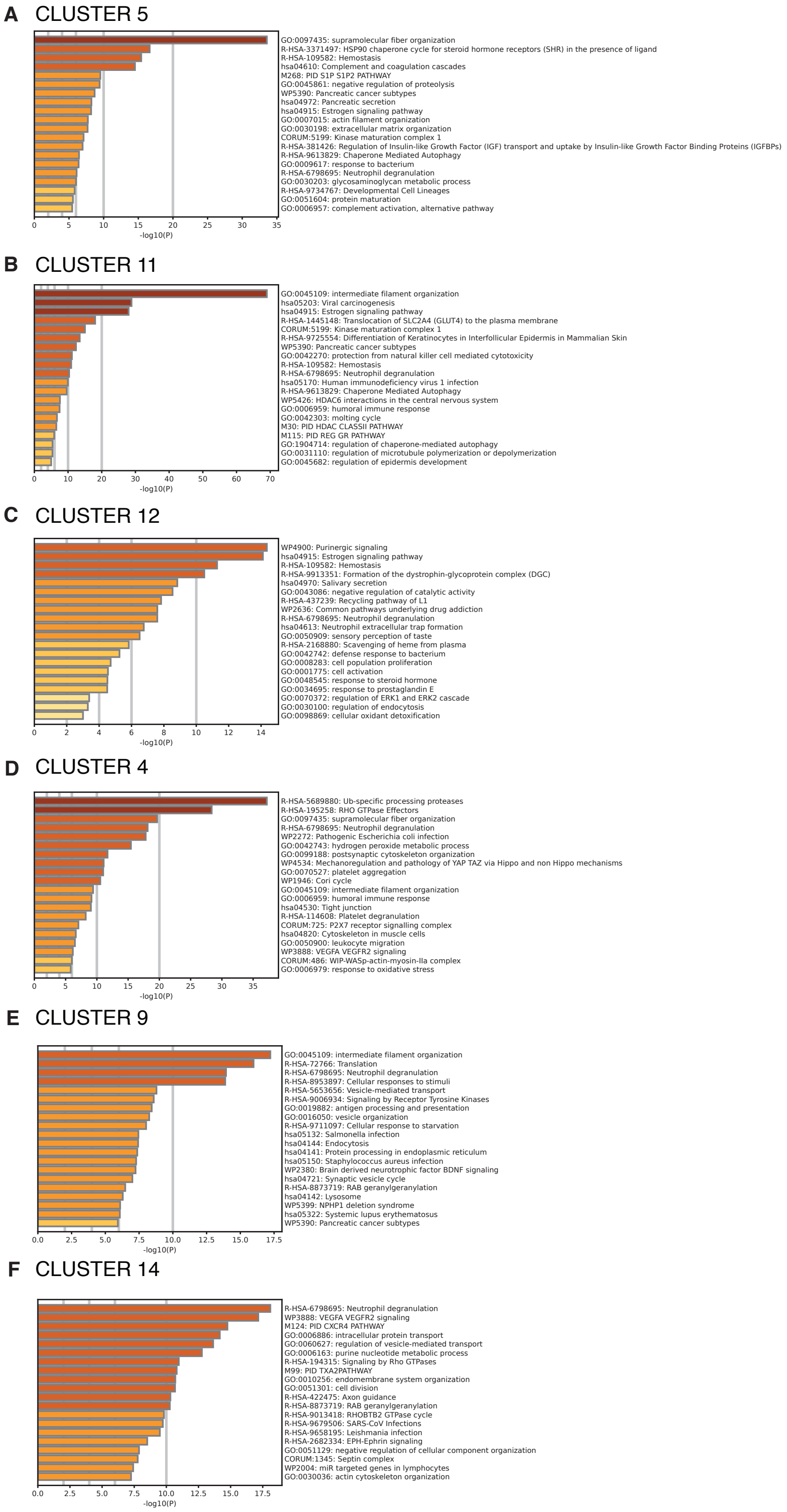


**Supplementary figure 8. Metascape analysis of proteins enriched in clusters of interest.** The metascape analysis reveals the functional enrichment of the associated proteins in the k-means clusters **A)** 5, **B)** 11, **C)** 12, **D)** 4, **E)** 9 and **F)** 14.

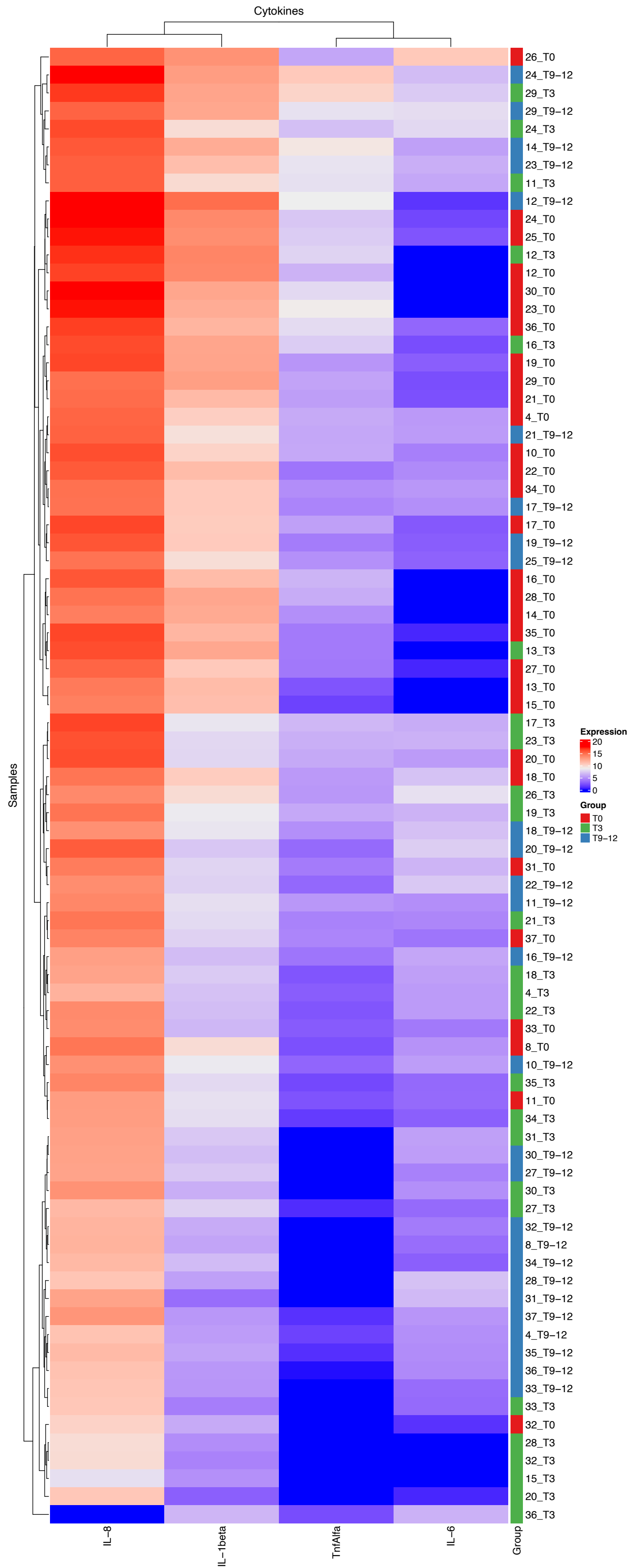


**Supplementary figure 9. Heatmap of cytokine expression (Log2) profiles.** In the clustering, samples collected from pwCFs at 0 months are shown in red, at 3 months in green, at 9-12 months in blue.
